# Health impact and cost of COVID-19 prophylaxis with monoclonal antibodies

**DOI:** 10.1101/2021.08.31.21262923

**Authors:** Abraham D. Flaxman, Rodal Issema, Ruanne V. Barnabas, Jennifer M. Ross

**Affiliations:** Institute for Health Metrics and Evaluation, University of Washington, Seattle, WA; Department of Epidemiology, University of Washington, Seattle, WA; International Clinical Research Center, Department of Global Health, University of Washington, Seattle, WA; Division of Allergy and Infectious Diseases, Department of Medicine, University of Washington, Seattle, WA

## Abstract

**Background:** The COVID-19 pandemic has led to over 600,000 deaths in the United States and continues to disrupt lives even as effective vaccines are available. We aimed to estimate the impact and health system cost of implementing post-exposure prophylaxis against household exposure to COVID-19 with monoclonal antibodies.

**Methods:** We developed a decision-analytical model analysis of results from a recent randomized controlled trial with complementary data on household demographic structure, vaccine coverage, and COVID-19 confirmed case counts for the representative month of May, 2021. The model population includes individuals of all ages in the United States by sex and race/ethnicity.

**Results:** In a month of similar intensity to May, 2021, in the USA, a monoclonal antibody post-exposure prophylaxis program reaching 50% of exposed unvaccinated household members aged 50+, would avert 1,813 (1,171 – 2,456) symptomatic infections, 526 (343 - 716) hospitalizations, and 83 (56 - 116) deaths. Assuming the unit cost of administering the intervention was US$ 1,264, this program would save the health system US$ 3,055,202 (−14,034,632 - 18,787,692).

**Conclusions:** Currently in the United States, health system and public health actors have an opportunity to improve health and reduce costs through COVID-19 post-exposure prophylaxis with monoclonal antibodies.

## Introduction

The coronavirus disease 2019 (COVID-19) pandemic, caused by severe acute respiratory syndrome coronavirus 2 (SARS-CoV-2), has led to more than 600,000 deaths in the United States since February, 2020, and continues to disrupt lives even as effective vaccines are available and vaccine coverage increases.^1,2^ Initially, nonpharmaceutical interventions led to sustained declines in SARS-CoV-2 infections and COVID-19 deaths, but this was followed by multiple resurgences in cases that are thought to be due to the emergence of new, more transmissible SARS-CoV-2 variants of concern.^3,4^ Over time, prevention and treatment interventions for SARS-CoV-2 have expanded dramatically from repurposed antivirals to highly effective vaccines and promising monoclonal antibodies.^5–7^ Despite the tremendous success of COVID-19 vaccine development and initial distribution, the pace of vaccination slowed in the US, with a sizeable proportion of eligible persons remaining unvaccinated.^8^ Thus, observed COVID-19 cases, hospitalizations, and deaths have risen again due to multiple causes including vaccination coverage below the threshold for herd immunity, new variants causing vaccine breakthrough infections, and potential declining immunity over time.^9^

In this setting of ongoing SARS-CoV-2 transmission, monoclonal antibody (mAb) therapies are an additional tool with demonstrated efficacy to prevent infection among unvaccinated individuals with a high-risk exposure to someone with SARS-CoV-2 infection.^10,11^ Antibody therapies are fast acting, since their ready-made antibodies can bind to antigen immediately, in contrast to vaccines that stimulate the body to produce an immune response over weeks.^12^ This property of fast-acting, time-limited protection makes antibody therapies potentially attractive for use in household exposure situations where unvaccinated household contacts are at high risk of acquiring infection over a short time interval and can be identified rapidly.^13^ However, antibodies are relatively costly to produce, which makes it important to assess their optimal use in health economic analyses.

Recently, a randomized, placebo-control trial of the antibody “cocktail” of casirivimab with imdevimab (cas/imdev; formerly REGN-COV2) demonstrated efficacy in preventing symptomatic COVID-19 and PCR-positive SARS-CoV-2 infection (asymptomatic or symptomatic) when given to asymptomatic, SARS-CoV-2 negative household members of people with COVID-19 within 96 hours of their household member testing positive, supporting an emergency use authorization for use as post-exposure prophylaxis among people who are unvaccinated or unlikely to mount a protective response following vaccination.^10,14^ Similarly, in a study of 966 nursing home residents and staff, prophylaxis with the monoclonal antibody bamlanivimab reduced symptomatic COVID-19 by >40%.^11^ Importantly, current evidence suggests that monoclonal antibody combinations continue to offer protection against new variants.^15,16^

In this decision-analytical analysis, we quantified the health impact and cost of a hypothetical post-exposure prophylaxis program where unvaccinated household members who have been exposed to COVID-19 are given monoclonal antibodies (mAb PEP), to understand the potential public-health significance of the approach recently shown to have strong clinical benefit.

## Methods

### Study design

We used a decision-analytical model to combine results from the cas/imdev randomized controlled trial with population data on household demographic structure, COVID-19 confirmed case counts and demographics, and vaccination coverage to estimate the number of symptomatic infections, hospitalizations, deaths, and health system cost for mAb PEP programs of varying intensities.^10,17^ Our focus was on the general population of the United States to provide actionable evidence for public health policy. The key decision points in our model are the coverage level and age targeting of the intervention.

The baseline strategy in our analysis was to not implement mAb PEP, and we compared this to implementing mAb PEP at 4 intensities of coverage, where 25%, 50%, 75%, or 100% of individuals with household exposure to someone with confirmed COVID-19 and age above the minimum age threshold received mAb PEP. For each of coverage level, we explored the impact and cost of 7 different age-based inclusion criteria (no minimum age for PEP, age 20+, 40+, 50+, 60+, 70+, 80+). We used a time horizon corresponding to one wave of SARS-CoV-2 transmission, roughly one month in duration, to evaluate the costs and impact of the program. We used a health system perspective to quantify the total cost, which included both the cost of the mAb PEP intervention and the (offset) cost of COVID-19 hospitalizations. We did not use time-discounting for costs or outcomes because of the short time horizon.

### Data

Efficacy data come from the phase 3, placebo-controlled, household randomized trial of the monoclonal antibody cocktail casirivimab with indevimab administered as subcutaneous injections to asymptomatic, SARS-CoV-2 negative household contacts of people with confirmed COVID-19.^10^ The primary outcome of symptomatic SARS-CoV-2 infection developed in 59 of 752 (7.8%) placebo recipients and 11 of 753 (1.5%) mAb PEP recipients, indicating an 81% risk reduction. From this trial, we incorporated a household secondary attack rate of 7.8%, which is conservatively low relative to a recent systematic review.^18^

To estimate the public health significance of implementing mAb PEP widely, we combined the effect size and secondary attack rate from the randomized trial with data on the demographic structure of US households, data on recent confirmed cases of COVID-19, and COVID-19 vaccine coverage and clustering by household. We used the public-use microdata sample (PUMS) data from the American Community Survey (ACS) to group individuals into households by age group, sex, and race/ethnicity demographic strata and estimate the average number of individuals that would have a household exposure to COVID-19 from an index case in any demographic stratum.^17^ For example, to estimate the number of Black females aged 80+ who would be exposed by a Black male age 30-39 with COVID-19, we identified all households in the PUMS data with a Black male aged 30-39 and counted the number of Black females aged 80+ in each household. Then, we used the arithmetic mean of these counts as our estimate of the number of people exposed. We excluded all individuals living in group quarters. To quantify uncertainty, we used non-parametric bootstrap resampling of 100 households with replacement for an index case in each demographic stratum.^19^

To find the age-, sex-, and race/ethnicity-specific rates of COVID-19 detection, we used COVID-19 confirmed case data from CDC for the month of May, 2021.^20^ We calculated the fraction of cases in each demographic stratum using a complete-case analysis that dropped rows with missing data on age, sex, or race/ethnicity, and then scaled these fractions to match the total count of cases including those with missing demographic data. We used non-parametric bootstrap resampling to quantify uncertainty.^19^

We modeled household clustering of COVID-19 vaccination status using the Kaiser Family Foundation survey from June, 2021.^8^ In this nationally-representative survey of US adults, 77% of vaccinated respondents indicated that everyone in their household also was vaccinated against COVID-19, while 69% of unvaccinated respondents reported that everyone in their household was unvaccinated. We used Bayes law to derive the fraction of confirmed cases who are unvaccinated from the population coverage and real-world efficacy data and combined this with survey data on the percentage of unvaccinated people who live with others where everyone in the household is unvaccinated (appendix page 1).^21^

We modeled the cost of COVID-19 hospitalization as $73,300 based on analysis by FAIR Health using ICD-10 procedure codes.^22^ We estimated the unit cost of mAb PEP to be $1250 from the federal government purchase price for the monoclonal antibody bamlanivimab.^23^ We estimated the cost of services related to mAb PEP administration as $114 based on a time-driven activity-based cost estimate reported for outpatient subcutaneous administration of another medication adjusted to 2020 US$ (appendix page 3).^24^

### Analytical methods

To estimate the cost and impact of PEP, we used an analytical model summarized by a decision tree (Figure 1) with a choice node for mAb PEP followed by chance nodes for symptomatic infection, hospitalization, and death. We used demographic data from confirmed cases of COVID-19 to identify households where PEP would be indicated, and then used demographic data on household structure to identify the age, sex, and race of the household members who could receive mAb PEP. Details of the approach and parameter values are provided in the appendix. We used this approach to balance the complexity needed to capture the hypothesized differences between impact by race/ethnicity with the simplicity of a multiplicative model structure. To investigate this hypothesis, we calculated the rates of infections, hospitalizations, and deaths averted, all stratified by race/ethnicity.

**Figure 1:**
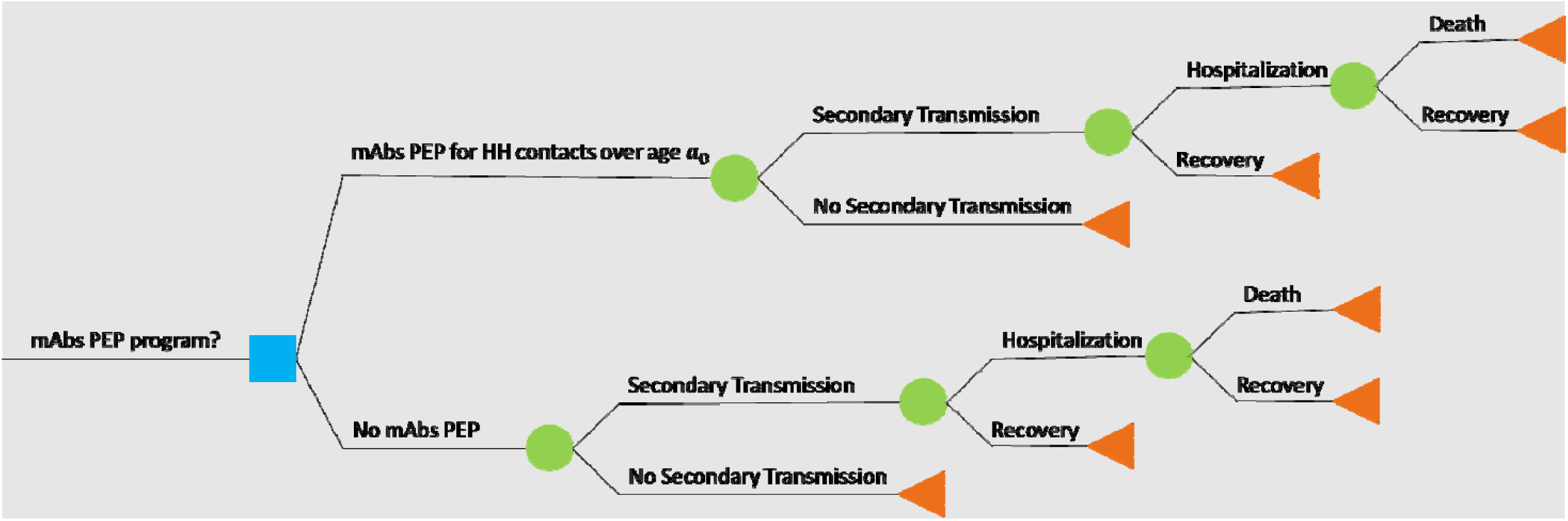
Decision tree representation of our analytical model, with a single choice node (blue square) for PEP for each individual household contact, followed by a series of chance nodes (green circles) for secondary infection, hospitalization, and mortality, leading eventually to terminal nodes (red triangles) for recovery or death.

### Model assumptions

The model assumes a sequential progression of COVID-19 severity, with infection sometimes progressing to hospitalization, which in turn sometimes progresses to death. With this approach, when stratified by age, sex, and race/ethnicity, the fraction of symptomatic infections progressing to hospitalization (modeled) is the same as the fraction of confirmed cases progressing to hospitalization (observed). The average cost of hospitalization was constant and did not vary by individual characteristics. Also, after stratifying by age group, sex, and race/ethnicity, the household structure of people with confirmed cases COVID-19 was assumed to match that of the general population.

We report estimates according to the CHEERS standards.^25^ We published a replication archive for the code used in this analysis.^26^

## Results

The cas/imdev PEP trial included 1555 participants (753 in treatment group and 752 in placebo), which we combined with confirmed cases data on 154,136 individuals, vaccine coverage survey data from 1,888 individuals, and household structure data derived from 3,190,040 individuals in 1,394,399 households. We estimate that the 154,136 confirmed cases of COVID-19 in May in the US resulted in at least 255,959 (95% UI 240,450-271,482) unvaccinated individuals with household exposure to COVID-19. Using the racial and ethnic characteristics of households and COVID-19 cases described above, we estimated unvaccinated household contacts to include 43,052 (16.8%) non-Hispanic Black individuals, 50,297 (19.7%) Hispanic individuals, 135,396 (52.9%) non-Hispanic white individuals, and 27,214 (10.6%) members of other non-Hispanic racial and ethnic groups.

A scenario providing PEP to 50% of unvaccinated household contacts age 50+ resulted in treatment of 28,187 (95% UI 26,336-30,536) individuals, with fewer individuals treated at higher age thresholds (Table 1). PEP coverage of 50% of unvaccinated contacts age 50+ averted 1,813 (95% UI 1,171 - 2,456) symptomatic COVID-19 cases, 526 (95% UI 343 - 716) hospitalizations, and prevented 83 (95% UI 56 - 116) deaths. Expanding the age threshold to 20+ years increased the averted burden to 5,056 (95% UI 3,277 - 6,990) symptomatic COVID-19 cases, 768 (95% UI 500 - 1,036) hospitalizations, and 93 (95% UI 61 - 129) deaths, while restricting to age 80+ years reduced the burden averted to 104 (95% UI 67 - 151) symptomatic COVID-19 cases, 67 (95% UI 44 - 97) hospitalizations, and 24 (95% UI 15 - 36) deaths for the same assumption of 50% coverage of unvaccinated contacts in each target age group. Expanding PEP coverage by half from 50% to 75% resulted in corresponding 50% reductions in symptomatic COVID-19 cases, hospitalizations, and deaths.

**Table 1:** mAb PEP treatments provided and COVID-19 outcomes predicted in age threshold scenarios

|  | **Household contacts** | **Unvaccinated household contacts** | **PEP coverage** | **Number Treated with PEP** | **Symptomatic COVID-19 from household exposure** | **Hospitalizations from household exposure** | **Deaths from household exposure** |
| --- | --- | --- | --- | --- | --- | --- | --- |
| Baseline (no PEP) | 381,499<br>(373,749 - 389,768) | 255,959<br>(240,450 - 271,482) | 0 | 0 | 20,075 (15,641-25,670) | 2,040 (1,595-2,572) | 227 (172-293) |
| **Age threshold for PEP** | **Household contacts in age group** | **Unvaccinated household contacts in age group** | **PEP coverage** | **Number Treated with PEP** | **Symptomatic COVID-19 from household exposure** | **Hospitalizations from household exposure** | **Deaths from household exposure** |
| ≥80 years | 4,799<br>(4,078 - 5,742) | 3,221 (2,711 - 4,015) | 50% | 1,611<br>(1,356-2,008) | 19,972 (15,562-25,540) | 1,972 (1,537-2,485) | 203 (155-261) |
| ≥50 years | 84,014<br>(80,635 - 88,132) | 56,374<br>(52,673 - 61,072) | 50% | 28,187<br>(26,336-30,536) | 18,262 (14,155-23,253) | 1,513 (1,170-1,861) | 143 (109-188) |
| ≥20 years | 234,447<br>(229,161 - 239,175) | 157,298<br>(148,345 - 167,114) | 50% | 78,649<br>(74,173-83,557) | 15,019 (11,607-18,855) | 1,271 (990-1,571) | 134 (102-176) |

The rates of averted COVID-19 cases, hospitalization, and death differed by race and ethnicity. Assuming 50% mAb PEP coverage of unvaccinated individuals age 50+, the estimated rate of averted secondary infections was 80, 44, 55, and 61 symptomatic infections per 10,000,000 person-months among non-Hispanic Black, Hispanic, non-Hispanic white, and non-Hispanic members of other racial and ethnic groups, respectively. The averted hospitalization rate per 10,000,000 person-months were 32, 15, 13, and 21, while the averted death rates per 10,000,000 person-months were 50, 31, 16, and 48 for the same races, respectively.

We estimate that without PEP, the cost of hospitalizations due to COVID-19 infections from household exposure would be 150 (95% UI 109-205) million dollars, while in a hypothetical scenario where 50% of unvaccinated household contacts age 50 or older receive PEP, the cost of hospitalizations would be 111 (95% UI 79-152) million dollars and the cost of PEP would be 36 (95% UI 30-44) million dollars, for a total of 147 (95% UI 113-191) million dollars, which is a savings of 3 (95% UI -14-19) million dollars compared to the without-PEP scenario. The corresponding costs in 80+ age scenario were 145 (95% UI 106-198) million dollars for hospitalizations and 2 (95% UI 2-3) million dollars for PEPE, for a total of 147 (95% UI 108-200) million dollars, which is a savings of 3 (95% UI 1-5) million dollars compared to the without-PEP scenario. Alternatively, 50% mAb PEP coverage in the 20+ age scenario would cost 93 (95% UI 65-128) million dollars in hospitalizations and 99 (95% UI 82-123) million dollars for PEP, totaling 193 (95% UI 154-233) million dollars, which is 43 (95% UI 14-75) million dollars more than in the without-PEP scenario (Fig 2).

**Figure 2:**
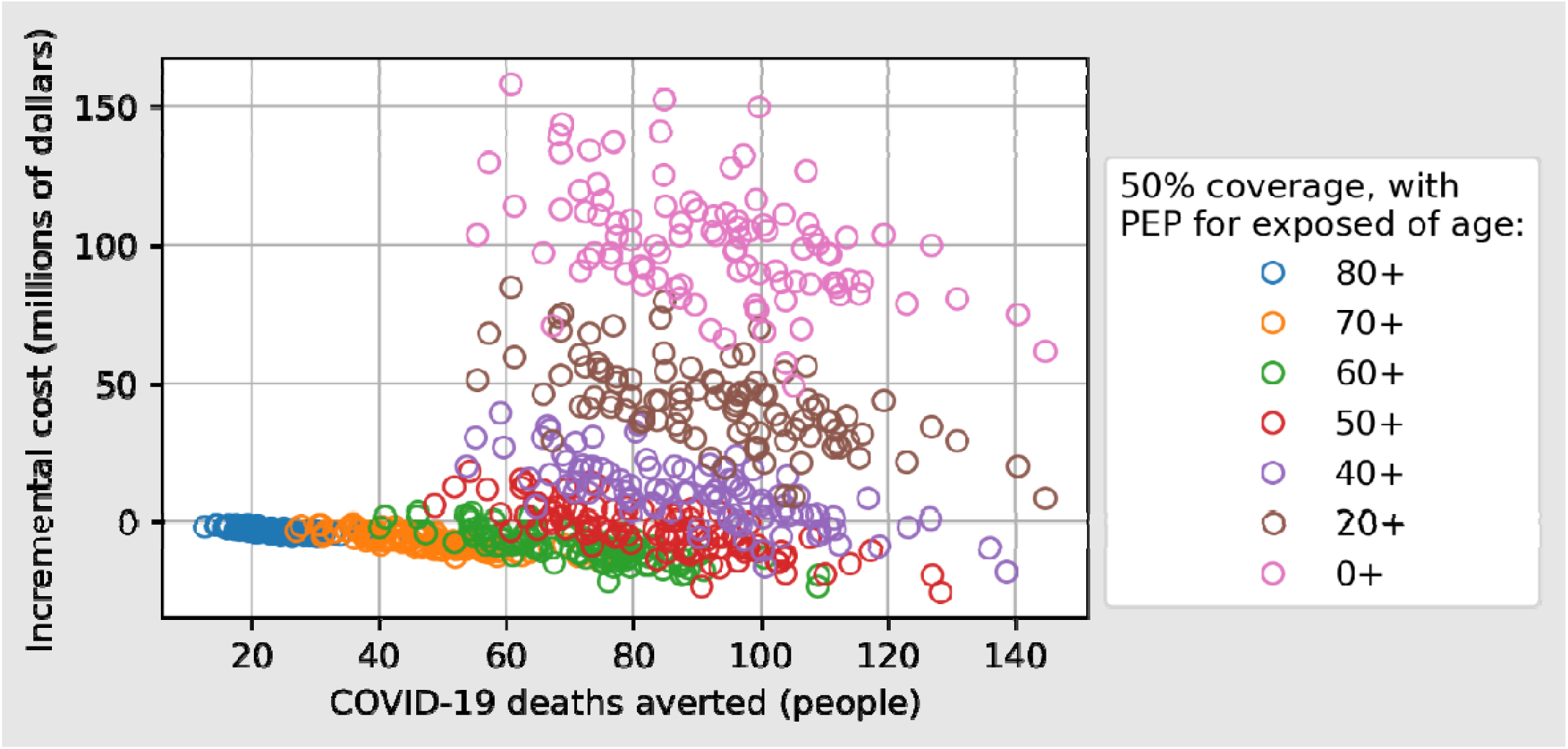
The number of COVID-19 deaths averted increases as the minimum age threshold for receiving PEP is decreased, and the incremental cost (including cost of PEP with mAbs plus cost of COVID-19 hospitalizations) shows the tradeoff between reducing hospitalization costs and increasing PEP costs, with a maximum reduction in incremental cost for a program that offers PEP to individuals aged 50 years or older.

## Discussion

As the COVID-19 pandemic continues in the US despite vaccination roll-out, we estimate that providing mAb PEP to unvaccinated household contacts of persons with COVID-19 age 50 or older would prevent COVID-19 disease and deaths and result in a small cost savings due to averted costs of hospitalization over one month of implementation in the US. Ours is a conservative lower bound of the estimate of the PEP benefit because we use a transmission probability for symptomatic COVID-19 of 8% observed in the imdev/casi mAb PEP trial, which is lower than the nearly 15% of household contacts who acquired SARS-CoV-2 infection with or without symptoms in household transmission studies, or the estimate of a 21.1% secondary attack rate within households in a recent meta-analysis.^18,27^ While persons who acquire SARS-CoV-2 infection and are asymptomatic do not contribute to health care costs, they can transmit infection to people who would develop symptoms and accrue health care costs. We do not incorporate COVID-19 cases and costs that occur from a subsequent round of transmission from household contacts to other individuals. Despite these conservative assumptions, the use of mAb PEP averts sufficient morbidity and mortality to be an efficient use of resources.

Acknowledging many unknowns about the future course of the COVID-19 pandemic, mAb PEP could have a beneficial role in COVID-19 combination prevention for several reasons. First, less than full vaccination coverage among US adults has left a persistent protection gap among people who could benefit from mAb PEP if exposed, including the same individuals who are more likely to share a household with other unvaccinated people at greatest risk for infection. Second, mAb PEP may be acceptable to an unvaccinated person with a high-risk household exposure because it offers rapid protection relative to initiating a multi-week vaccine series and could be packaged with follow-up vaccination. Third, as cases rise or with future shifts in dominant SARS-CoV-2 genotypes/variants, mAb PEP can be rapidly manufactured and combined if necessary to respond to new variants. Finally, beyond households, mAb PEP may be utilized in other settings with high-risk exposures, such as health care settings or long-term care facilities or among immunocompromised people who are less likely to mount an immune response to vaccination.^28^

Demonstrating the population health benefit and economic value of mAb PEP is crucial to overcoming the initial logistical challenges of implementing a novel intervention. To our knowledge, this is among the first health economic analysis of COVID-19 mAb PEP within the US population. Methodological strengths of our analysis include use of large, publicly available datasets on household composition, COVID-19 cases, and vaccine coverage; high-quality efficacy data observed in a randomized controlled trial; and incorporation of uncertainty measures throughout the model.

This analysis also has limitations. First, population structure may have changed due to housing insecurity, work disruption, and general adaptation during more than a year of social distancing in response to COVID-19 pandemic. Second, there was missing data in confirmed cases, especially about race, hospitalizations, and deaths. Third, we modeled the whole US population without differentiating regionally for vaccine coverage, household composition, and COVID-19 case activity. Fourth, our transmission probability parameter from the clinical trial does not incorporate increased infectiousness due to the delta variant, which is still being characterized in the household setting.^29^ Fifth, SARS-CoV-2 viral evolution causes challenges in the forecasting the future PEP benefit, as new SARS-CoV-2 variants may reduce mAb PEP efficacy. Some variants have already demonstrated resistance to neutralization by monoclonal antibodies, though early preclinical and human studies indicate protection by imdev/casi against the SARS-CoV-2 variants of concern including the delta variant.^15^ Finally, we assume that unvaccinated individuals are susceptible to SARS-CoV-2 infection without accounting for partial immunity from prior SARS-CoV-2 infection.

## Conclusion

In summary, currently in the United States, health system and public health actors have an opportunity to improve health and reduce costs through COVID-19 post-exposure prophylaxis with monoclonal antibodies.

## Data Availability

The analysis used publicly-available data. The code for the analysis is published in a replication archive at https://doi.org/10.5281/zenodo.5338919.

## Funding

This work was supported by funding from the National Science Foundation (award DMS-1839116).

## Competing interests

ADF has consulted recently for Janssen; SwissRe; Sanofi; Merck for Mothers; and Agathos, Ltd. Other authors have no competing interested to disclose.

## Ethical review

These research activities used no identifiable private information and were therefore exempt from institutional board review.

