## Supplemental appendix for "Health impact and cost of COVID-19 prophylaxis with monoclonal antibodies"

### Supplemental Methods

#### Analytical steps:

1. Estimate the number of COVID-19 case count, $n_{1}$, and the age-/sex-/race-stratified case counts as

$$n_{1}\left( a,s,r \right)=\# cases for strata \left( a,s,r \right).$$

2. Estimate the stratified number of unvaccinated household contacts who might benefit from mAbs PEP as

$$n_{2}\left( a,s,r \right)=\sum_{a^{'},s^{'},r^{'}} p_{\mathrm{unvax}}\times n_{1}(a^{'},s^{'},r^{'})\times n_{hh}^{a^{'},s^{'},r^{'}}\left( a,s,r \right),$$

where $n_{hh}^{a^{'},s^{'},r^{'}}\left( a,s,r \right)$ denotes the mean number of *other* people in strata $\left( a,s,r \right)$ who live in a household with a person in strata $\left( a',s',r' \right)$ and $p_{\mathrm{unvax}}$ denotes the probability that a COVID-19 case occurs in a completely unvaccinated household.

3. Calculate the probability that a COVID-19 case occurs in a completely unvaccinated household as:

- Equation 1:

Pr(CC)=(Pr(CC|vax)×Pr(vax))+(Pr(CC|unvax)×Pr(unvax))

- Equation 2:

Pr(unvax|CC)=Pr(CC|unvax)×Pr(unvax)Pr(CC)

- Equation 3:

Pr(CC|vax)=(1−efficacy)×Pr(CC|unvax)

- Substitute Equation 3 into Equation 1:

Pr(CC)=((1−efficacy)×Pr(CC|unvax)×Pr(vax))+(Pr(CC|unvax)×Pr(unvax))

=Pr(CC|unvax)(((1−efficacy) ×Pr(vax))+Pr(unvax))*=*

- Substitute Equation 1 into Equation 2:

Pr(unvax|CC)=Pr(CC|unvax)×Pr(unvax)Pr(CC|unvax)(((1−efficacy) ×Pr(vax))+Pr(unvax))*P*

=Pr(unvax)((1−efficacy) ×Pr(vax))+Pr(unvax)

4. Estimate the number of people who receive PEP in scenarios with a range of coverage levels $c$ and minimum age $a_{0}$ for receiving PEP as

$$n_{\mathrm{PEP}}^{c,a_{0}}\left( a,s,r \right)=c\times n_{2}\left( a,s,r \right)\times[a\geq a_{0}],$$

where $[a\geq a_{0}]$ is equal to 1 if $a\geq a_{0}$ and 0 otherwise.

5. Estimate the number of people who develop a symptomatic COVID-19 infection in each scenario as

$$n_{\mathrm{COVID}}^{c,a_{0}}\left( a,s,r \right)=\mathrm{ar}_{\bar{\mathrm{PEP}}}\times\left( n_{2}\left( a,s,r \right)-n_{\mathrm{PEP}}^{c,a_{0}}\left( a,s,r \right) \right)+\mathrm{ar}_{\mathrm{PEP}}{\times n}_{\mathrm{PEP}}^{c,a_{0}}\left( a,s,r \right),$$

where $\mathrm{ar}_{\bar{\mathrm{PEP}}}$ is the secondary attack rate without mAbs and $\mathrm{ar}_{\mathrm{PEP}}$ is the secondary attack rate with mAbs.

6. Estimate the number of hospitalizations in each scenario as

$$n_{\mathrm{hosp}}^{c,a_{0}}\left( a,s,r \right)=\mathrm{hr}\left( a,s,r \right)\times n_{\mathrm{COVID}}^{c,a_{0}}\left( a,s,r \right),$$

where $\mathrm{hr}\left( a,s,r \right)$ is the hospitalization rate for strata $\left( a,s,r \right)$.

7. Estimate the number of deaths in each scenario as

$$n_{\mathrm{death}}^{c,a_{0}}\left( a,s,r \right)=\mathrm{hfr}\left( a,s,r \right)\times n_{\mathrm{hosp}}^{c,a_{0}}\left( a,s,r \right),$$

where $\mathrm{hfr}\left( a,s,r \right)$ is the hospitalization fatality ratio for strata $\left( a,s,r \right)$.

8. Estimate the cost of administering mAbs and the cost of COVID-19 hospitalizations as

$$c_{\mathrm{mAbs}}^{c,a_{0}}=uc_{\mathrm{mAbs}}\times\sum_{a,s,r} n_{\mathrm{PEP}}^{c,a_{0}}(a,s,r),$$

$$c_{\mathrm{hosp}}^{c,a_{0}}=uc_{\mathrm{hosp}}\times\sum_{a,s,r} n_{\mathrm{hosp}}^{c,a_{0}}(a,s,r),$$

where $uc_{\mathrm{mAbs}}$ and $uc_{\mathrm{hosp}}$ are the unit costs for mAbs and COVID-19 hospitalization.

#### Parameters, values, data sources, and analytical methods:

- $n_{1}\left( a,s,r \right)$ comes from CDC confirmed cases; to deal with missing data, we computed the fraction of cases by age, sex, and race/ethnicity from the rows with complete data (complete-case analysis) [[and then scaled this value to the total number of cases detected during May 2021.]]. We selected age groups to match those available in the CDC data, and collapsed race/ethnicity to white, Black, Hispanic, and all other race/ethnicities.^1^
- $n_{hh}^{a^{'},s^{'},r^{'}}\left( a,s,r \right)$ comes from ACS PUMS data, which includes age in years, sex, race/ethnicity, and household ID.^2^
- We examined scenarios corresponding to all combinations of the age thresholds matched to the age groups from CDC data, and coverage levels of 0, 33%, 50%, 67%, and 100%.
- $\mathrm{ar}_{\bar{\mathrm{PEP}}}=0.078$ and $\mathrm{ar}_{\mathrm{PEP}}=0.014$ came from the imdev/casi RCT.^3^
- We calculated $\mathrm{hr}\left( a,s,r \right)$ from CDC data. We dropped all cases where age, sex, race/ethnicity, or hospitalization status was unknown.^1^
- We calculated $\mathrm{hfr}\left( a,s,r \right)$ similarly from CDC data, by dropping all cases which were not hospitalized or where age, sex, race/ethnicity, or mortality status was unknown.
- We determined the unit costs $uc_{\mathrm{mAbs}}=\$1,264$ using $1,250 as the cost of the medication and $114 as the cost of administration.^4^ We calculated the cost of administration as $111 using the subcutaneous administration scenario of a time-driven activity-based cost estimate by Wallace, et al, and adjusted to 2020 US$ using the Bureau of Labor Statistics Consumer Price Index calculator.^5^ We estimated the unit cost of hospitalization $uc_{\mathrm{hosp}}=\$73,300$ from recent literature.^6^

### Reporting checklist for economic evaluation of health interventions (CHEERS)

|  |  | Reporting Item | Page Number |
| --- | --- | --- | --- |
| **Title** |  |  |  |
|  | [#1](https://www.goodreports.org/reporting-checklists/cheers/info/#1) | Identify the study as an economic evaluation or use more specific terms such as “cost-effectiveness analysis”, and describe the interventions compared. | 1 |
| **Abstract** |  |  |  |
|  | [#2](https://www.goodreports.org/reporting-checklists/cheers/info/#2) | Provide a structured summary of objectives, perspective, setting, methods (including study design and inputs), results (including base case and uncertainty analyses), and conclusions | 2 |
| **Introduction** |  |  |  |
| Background and objectives | [#3](https://www.goodreports.org/reporting-checklists/cheers/info/#3) | Provide an explicit statement of the broader context for the study. Present the study question and its relevance for health policy or practice decisions | 3-4 |
| **Methods** |  |  |  |
| Target population and subgroups | [#4](https://www.goodreports.org/reporting-checklists/cheers/info/#4) | Describe characteristics of the base case population and subgroups analysed, including why they were chosen. | 5 |
| Setting and location | [#5](https://www.goodreports.org/reporting-checklists/cheers/info/#5) | State relevant aspects of the system(s) in which the decision(s) need(s) to be made. | 5 |
| Study perspective | [#6](https://www.goodreports.org/reporting-checklists/cheers/info/#6) | Describe the perspective of the study and relate this to the costs being evaluated. | 5 |
| Comparators | [#7](https://www.goodreports.org/reporting-checklists/cheers/info/#7) | Describe the interventions or strategies being compared and state why they were chosen. | 5 |
| Time horizon | [#8](https://www.goodreports.org/reporting-checklists/cheers/info/#8) | State the time horizon(s) over which costs and consequences are being evaluated and say why appropriate. | 5 |
| Discount rate | [#9](https://www.goodreports.org/reporting-checklists/cheers/info/#9) | Report the choice of discount rate(s) used for costs and outcomes and say why appropriate | 5 |
| Choice of health outcomes | [#10](https://www.goodreports.org/reporting-checklists/cheers/info/#10) | Describe what outcomes were used as the measure(s) of benefit in the evaluation and their relevance for the type of analysis performed | 5 |
| Measurement of effectiveness | [#11a](https://www.goodreports.org/reporting-checklists/cheers/info/#11a) | Single study-based estimates: Describe fully the design features of the single effectiveness study and why the single study was a sufficient source of clinical effectiveness data | 5 |
| Measurement of effectiveness | [#11b](https://www.goodreports.org/reporting-checklists/cheers/info/#11b) | Synthesis-based estimates: Describe fully the methods used for identification of included studies and synthesis of clinical effectiveness data | NA |
| Measurement and valuation of preference based outcomes | [#12](https://www.goodreports.org/reporting-checklists/cheers/info/#12) | If applicable, describe the population and methods used to elicit preferences for outcomes. | NA |
| **Estimating resources |  |  |  |
| and costs ** |  |  |  |
|  | [#13a](https://www.goodreports.org/reporting-checklists/cheers/info/#13a) | Single study-based economic evaluation: Describe approaches used to estimate resource use associated with the alternative interventions. Describe primary or secondary research methods for valuing each resource item in terms of its unit cost. Describe any adjustments made to approximate to opportunity costs | NA |
| **Methods** |  |  |  |
| Estimating resources and costs | [#13b](https://www.goodreports.org/reporting-checklists/cheers/info/#13b) | Model-based economic evaluation: Describe approaches and data sources used to estimate resource use associated with model health states. Describe primary or secondary research methods for valuing each resource item in terms of its unit cost. Describe any adjustments made to approximate to opportunity costs. | 7 |
| Currency, price date, and conversion | [#14](https://www.goodreports.org/reporting-checklists/cheers/info/#14) | Report the dates of the estimated resource quantities and unit costs. Describe methods for adjusting estimated unit costs to the year of reported costs if necessary. Describe methods for converting costs into a common currency base and the exchange rate. | 7 |
| Choice of model | [#15](https://www.goodreports.org/reporting-checklists/cheers/info/#15) | Describe and give reasons for the specific type of decision analytical model used. Providing a figure to show model structure is strongly recommended. | 8 |
| Assumptions | [#16](https://www.goodreports.org/reporting-checklists/cheers/info/#16) | Describe all structural or other assumptions underpinning the decision-analytical model. | 8 |
| Analytical methods | [#17](https://www.goodreports.org/reporting-checklists/cheers/info/#17) | Describe all analytical methods supporting the evaluation. This could include methods for dealing with skewed, missing, or censored data; extrapolation methods; methods for pooling data; approaches to validate or make adjustments (such as half cycle corrections) to a model; and methods for handling population heterogeneity and uncertainty. | 6 |
| **Results** |  |  |  |
| Study parameters | [#18](https://www.goodreports.org/reporting-checklists/cheers/info/#18) | Report the values, ranges, references, and, if used, probability distributions for all parameters. Report reasons or sources for distributions used to represent uncertainty where appropriate. Providing a table to show the input values is strongly recommended. | Supplement |
| Incremental costs and outcomes | [#19](https://www.goodreports.org/reporting-checklists/cheers/info/#19) | For each intervention, report mean values for the main categories of estimated costs and outcomes of interest, as well as mean differences between the comparator groups. If applicable, report incremental cost-effectiveness ratios. | 11 |
| Characterising uncertainty | [#20a](https://www.goodreports.org/reporting-checklists/cheers/info/#20a) | Single study-based economic evaluation: Describe the effects of sampling uncertainty for the estimated incremental cost and incremental effectiveness parameters, together with the impact of methodological assumptions (such as discount rate, study perspective). | NA |
| Characterising uncertainty | [#20b](https://www.goodreports.org/reporting-checklists/cheers/info/#20b) | Model-based economic evaluation: Describe the effects on the results of uncertainty for all input parameters, and uncertainty related to the structure of the model and assumptions. | 6 |
| Characterising heterogeneity | [#21](https://www.goodreports.org/reporting-checklists/cheers/info/#21) | If applicable, report differences in costs, outcomes, or cost effectiveness that can be explained by variations between subgroups of patients with different baseline characteristics or other observed variability in effects that are not reducible by more information. | NA |
| **Discussion** |  |  |  |
| Study findings, limitations, generalisability, and current knowledge | [#22](https://www.goodreports.org/reporting-checklists/cheers/info/#22) | Summarise key study findings and describe how they support the conclusions reached. Discuss limitations and the generalisability of the findings and how the findings fit with current knowledge. | 12-13 |
| **Other** |  |  |  |
| Source of funding | [#23](https://www.goodreports.org/reporting-checklists/cheers/info/#23) | Describe how the study was funded and the role of the funder in the identification, design, conduct, and reporting of the analysis. Describe other non-monetary sources of support | 14 |
| Conflict of interest | [#24](https://www.goodreports.org/reporting-checklists/cheers/info/#24) | Describe any potential for conflict of interest of study contributors in accordance with journal policy. In the absence of a journal policy, we recommend authors comply with International Committee of Medical Journal Editors recommendations | 14 |

None The CHEERS checklist is distributed under the terms of the Creative Commons Attribution License CC-BY-NC. This checklist can be completed online using <https://www.goodreports.org/>, a tool made by the [EQUATOR Network](https://www.equator-network.org) in collaboration with [Penelope.ai](https://www.penelope.ai)
